# How is Exercise Prescribed for Hemodialysis Patients? A Scoping Review Protocol

**DOI:** 10.1101/2021.12.21.21268178

**Authors:** Heitor S. Ribeiro, Francini P. Andrade, Diogo V. Leal, Juliana S. Oliveira, Kenneth R. Wilund, João L. Viana

**Author notes:** Corresponding author: João L. Viana, PhD, Research Center in Sports Sciences, Health Sciences and Human Development – CIDESD, University of Maia – ISMAI, Av. Carlos Oliveira Campos - Castelo da Maia, Postal Code: 4475-690, Maia, Portugal. Twitter: @viana_JL.

## Abstract

**Objective:** The objective of this scoping review is to describe how exercise has been prescribed for hemodialysis patients.

**Introduction:** Exercise interventions have received more attention from the nephrology community in the last few years. Despite some limitations in the findings, there is currently robust evidence suggesting that exercise is clinically important and provides benefits to hemodialysis patients. Even so, there is little evidence precisely detailing and describing how exercise can be prescribed and delivered for this population.

**Inclusion criteria:** Based on the PCC framework, we will review and include evidence from hemodialysis patients (**P**articipants); describing exercise interventions (**C**oncept); in all settings and designs (**C**ontext). The evidence that included any other kidney replacement therapy other than hemodialysis will be excluded.

**Methods:** This review will follow the JBI methodology for scoping reviews and the PRISMA-ScR. We will perform a comprehensive literature search using MEDLINE, EMBASE, SPORTDiscuss, CINAHL, and LILACS databases without date or language restrictions from inception until December 2021. Websites, books, and guidelines from prominent societies and associations will also be searched. Experimental, quasi-experimental, observational, and protocol evidence from adults with chronic kidney disease (≥18 years) undergoing hemodialysis that prescribed exercise as an intervention will be considered. Two independent reviewers will screen title and abstract and perform the full-text review. Data extraction will be done by the main reviewer and checked by a second reviewer. Data characterizing the exercise interventions (*e.g*., type, setting, frequency, duration, intensity, volume, progression, periodization, professionals involved, etc.) will be extracted from selected evidence. The qualitative and quantitative results will be synthesized and presented in tables and figures along with a narrative summary.

## Introduction

Adults with chronic kidney disease (CKD) undergoing maintenance hemodialysis (HD) treatment usually have highly sedentary behaviour^1,2^, leading to musculoskeletal impairments and an increased prevalence of sarcopenia and frailty^3,4^. Exercise programs (*e.g*., resistance/strength, aerobic, and combined) have been proposed as therapeutic interventions to mitigate these outcomes^5,6^. Previous evidence have also shown that exercise programs produce positive effects on cardiorespiratory fitness, physical function, quality of life, neuropsychological disorders, and sleep quality in hemodialysis patients^7–9^.

While exercise interventions have received much greater attention from the nephrology community in the last few years^10^, there is still little evidence showing the implementation of exercise as part of the standard care for hemodialysis patients^11^. This indicates that apart from better understanding the benefits of prescribing exercise, the nephrology community requires more pragmatic and detailed intervention protocols that describe how to deliver and implement sustainable exercise programs for this population.

A preliminary search of MEDLINE, the Cochrane Database of Systematic Reviews and Joanna Briggs Institute (JBI) Evidence Synthesis was conducted. While systematic reviews on the effects of exercise on hemodialysis patients are ongoing, no scoping reviews (published or in progress) mapping the characteristics of exercise prescription for these patients were identified. Thus, the objective of this scoping review is to describe how exercise has been prescribed for hemodialysis patients.

## Review question

I. What types of exercise interventions are most frequently delivered to adults with CKD undergoing maintenance hemodialysis?
II. What are the main characteristics and variables adopted to prescribe exercise interventions to adults with CKD undergoing hemodialysis?

## Eligibility criteria

### Participants

This scoping review will consider evidence from reports/studies that prescribed exercise interventions for adults with CKD (≥18 years) from all genders (*i.e*., female, male, non-binary, etc.) undergoing hemodialysis. Evidence that included other kidney replacement therapies (*e.g*., peritoneal dialysis and kidney transplant), non-dialysis, pediatric patients (<18 years), or acute kidney injury will not be considered.

### Concept

We will consider any evidence from reports/studies that prescribed or recommended exercise interventions for adults with CKD undergoing maintenance hemodialysis. Exercise is classically defined as a “physical activity that is planned, structured, repetitive, and purposive in the sense that improvement or maintenance of one or more components of physical fitness and health is an objective”^12^. Non-experimental evidence, such as websites, books, and guidelines from prominent societies and associations that recommended or described exercise interventions will also be considered. We will consider all exercise possibilities and types that have been prescribed and recommended worldwide.

### Context

No restriction will be applied to the setting or design. Evidence from all available sources regarding exercise prescription during dialysis, out of dialysis, in a fitness center, home-based, in-hospital, or others will be considered.

### Types of Sources

This scoping review will consider all study designs and evidence (*e.g*., experimental, quasi-experimental, observational, protocol, etc.). In addition, grey literature sources, websites, books, and guidelines from prominent societies and associations will also be searched. On the other hand, systematic reviews, conference abstracts, commentaries, and letters to the editor will not be considered.

## Methods

The proposed scoping review will be conducted in accordance with the JBI methodology for scoping reviews^13^ and the Preferred Reporting Items for Systematic Reviews and Meta-Analyses extension for Scoping Reviews (PRISMA-ScR)^14^.

### Search strategy

The search strategy will aim to locate both peer-review published studies and non-scientific evidence. An initial limited search of MEDLINE was undertaken to identify articles on the topic. The text words contained in the titles and abstracts of relevant articles, and the index terms used to describe the articles were used to develop a full search strategy for MEDLINE, EMBASE, SPORTDiscuss, CINAHL, and LILACS databases (see Appendix 1). The search strategy, including all identified keywords and index terms, will be adapted for each included database and/or information source. Databases will be searched from the date of inception to December 2021. The reference list of all included sources of evidence will be screened for additional studies. We will not apply language or date restrictions. Sources of unpublished studies and grey literature to be searched include Google Scholar and Google.com.

### Source of Evidence selection

Following the search, all identified citations will be collated and uploaded into Covidence software (Veritas Health Innovation, Melbourne, AU) and duplicates removed for the screening process. Following a pilot test, titles and abstracts will then be screened by two independent reviewers (H.S.R and F.P.A) for assessment against the inclusion criteria for the review. Potentially relevant sources will be retrieved in full. The full text of selected evidence will be assessed in detail against the inclusion criteria by the same two independent reviewers. Reasons for exclusion of sources of evidence at full text that do not meet the inclusion criteria will be recorded and reported in the scoping review. Any disagreements that arise between the reviewers at each stage of the selection process will be resolved through discussion, or with an additional reviewer (D.V.L). The results of the search and the study inclusion process will be reported in full in the final scoping review and presented in PRISMA-ScR flow diagram^14^.

### Data Extraction

Data will be extracted from evidence included in the scoping review by the main reviewer (H.S.R) and checked for a second reviewer (F.P.A) using a data extraction spreadsheet developed by the reviewers adapted from JBI Manual for Evidence Synthesis^15^. The data extracted will include specific details about how exercise has been prescribed for adults with CKD undergoing maintenance hemodialysis (*e.g*., type, setting, frequency, duration, intensity, volume, progression, periodization, professionals involved, etc.). Also, additional general information will be extracted, such as author(s), year of publication, source of evidence, and country. A draft extraction form is provided (see Appendix 2). The draft data extraction tool will be modified and revised as necessary during the process of extracting data from each included evidence source. Any disagreements that arise between the reviewers will be resolved through discussion, or with an additional reviewer (D.V.L). If necessary, authors of papers will be contacted to request missing or additional data.

### Data Analysis and Presentation

The qualitative and quantitative results will be synthesized and presented in tables and figures along with a narrative summary. The frequencies of the extracted data will be displayed in network diagrams and/or heat maps. A narrative summary will accompany the illustrations and will describe how the results relate to the reviews objective and questions. The findings from the scoping review will be reported in accordance with PRISMA-ScR guideline^14^.

## Data Availability

All data produced in the present work are contained in the manuscript.

## Acknowledgements and Funding

We acknowledge the Research Center in Sports Sciences, Health Sciences and Human Development, CIDESD, supported by the Portuguese Foundation of Science and Technology (UID/04045/2020).

### Appendices

#### Appendix I: Search strategy

##### MEDLINE – 5,369 references (Dated 17 December 2021)

1. exp renal insufficiency, chronic/ or exp kidney diseases/
2. end-stage renal or end-stage kidney or endstage renal or endstage kidney or chronic kidney or chronic renal
3. ESRF or ESKF or ESRD or ESKD or CKF or CKD or CRF or CRD
4. exp renal replacement therapy/ or exp renal dialysis
5. dialysis or h?emodialysis or h?emofiltration or h?emodiafiltration
6. or/1-5
7. exp exercise/ or exp exercise therapy
8. (exercise or resistance training or strength training or aerobic training or endurance training or cycling training or combined exercise training or physical rehabilitation or physiotherapy or physical therapy).mp.
9. 9. or/7-8
10. 6 and 9
11. limit 10 to humans

##### Embase (OvidSP) – 15,988 references (Dated 20 December 2021)

1. exp renal insufficiency, chronic/ or exp kidney diseases/
2. ((kidney or renal) adj5 (disease* or injur* or insufficienc* or failure*)).mp
3. (end-stage renal or end-stage kidney or endstage renal or endstage kidney or chronic kidney or chronic renal).mp
4. (ESRF or ESKF or ESRD or ESKD or CKF or CKD or CRF or CRD).mp
5. exp renal replacement therapy/ or exp renal dialysis
6. (dialysis or h?emodialysis or h?emofiltration or h?emodiafiltration).mp
7. or/1-6
8. exp exercise/ or exp exercise therapy
9. (exercise or resistance training or strength training or aerobic training or endurance training or cycling training or combined exercise training or physical rehabilitation or physiotherapy or physical therapy).mp.
10. or/8-9
11. 7 and 10
12. limit 11 to human

##### CINAHL (EBSCO) – 972 references (Dated 20 December 2021)

1. (MH “Renal Insufficiency+”)
2. MH renal insufficienc* OR kidney insufficienc* OR renal diseas* OR kidney diseas*
3. MH chronic kidney disease OR chronic renal insufficiency
4. MH CKF OR CKD OR CRF OR CRD
5. S1 OR S2 OR S3 OR S4
6. MH exercise OR exercise physical
7. MH resistance training OR strength training OR aerobic training OR cycling training OR combined exercise training OR physical rehabilitation OR physiotherapy OR physical therapy
8. S6 OR S7
9. S5 AND S8

##### LILACS (BVS) – 11 references (Dated 20 December 2021)

(Renal Insufficiency OR Renal Insufficiency, Chronic OR Kidney Failure, Chronic OR renal replacement therapy OR Continuous Renal Replacement Therapy OR Hemofiltration OR Hemoperfusion OR Hybrid Renal Replacement Therapy OR Intermittent Renal Replacement Therapy OR Renal Dialysis) AND (exercise OR exercise physical OR resistance training OR strength training OR aerobic training OR cycling training OR combined exercise training OR physical rehabilitation OR physiotherapy OR physical therapy)

##### SPORTDiscuss (EBSCO) – 202 references (Dated 20 December 2021)

1. Hemodialysis
2. TI hemodialysis OR AB hemodialysis OR TI haemodialysis OR AB haemodialysis
3. TI renal insufficiency OR AB renal insufficiency
4. TI kidney failure OR AB kidney failure OR TI renal failure OR AB renal failure
5. TI renal replacement therapy OR AB renal replacement therapy OR TI kidney replacement therapy OR AB kidney replacement therapy
6. 6. 1-5 OR
7. Exercise
8. TI exercise OR AB exercise
9. TI “Physical conditioning” OR AB “Physical conditioning”
10. TI “Resistance training” OR AB “Resistance training”
11. TI “strength training” OR AB “strength training”
12. TI “Functional training” OR AB “Functional training”
13. TI “Aerobic activities” OR AB “Aerobic activities” OR TI “Aerobic activity” OR AB “Aerobic activity”
14. TI “Cardiovascular activities” OR AB “Cardiovascular activities” OR TI “Cardiovascular activity” OR AB “Cardiovascular activity”
15. TI “Endurance activities” OR AB “Endurance activities” OR TI “Endurance activity” OR AB “Endurance activity”
16. 7-15 OR
17. S6 AND S16

#### Appendix II: Data extraction tool

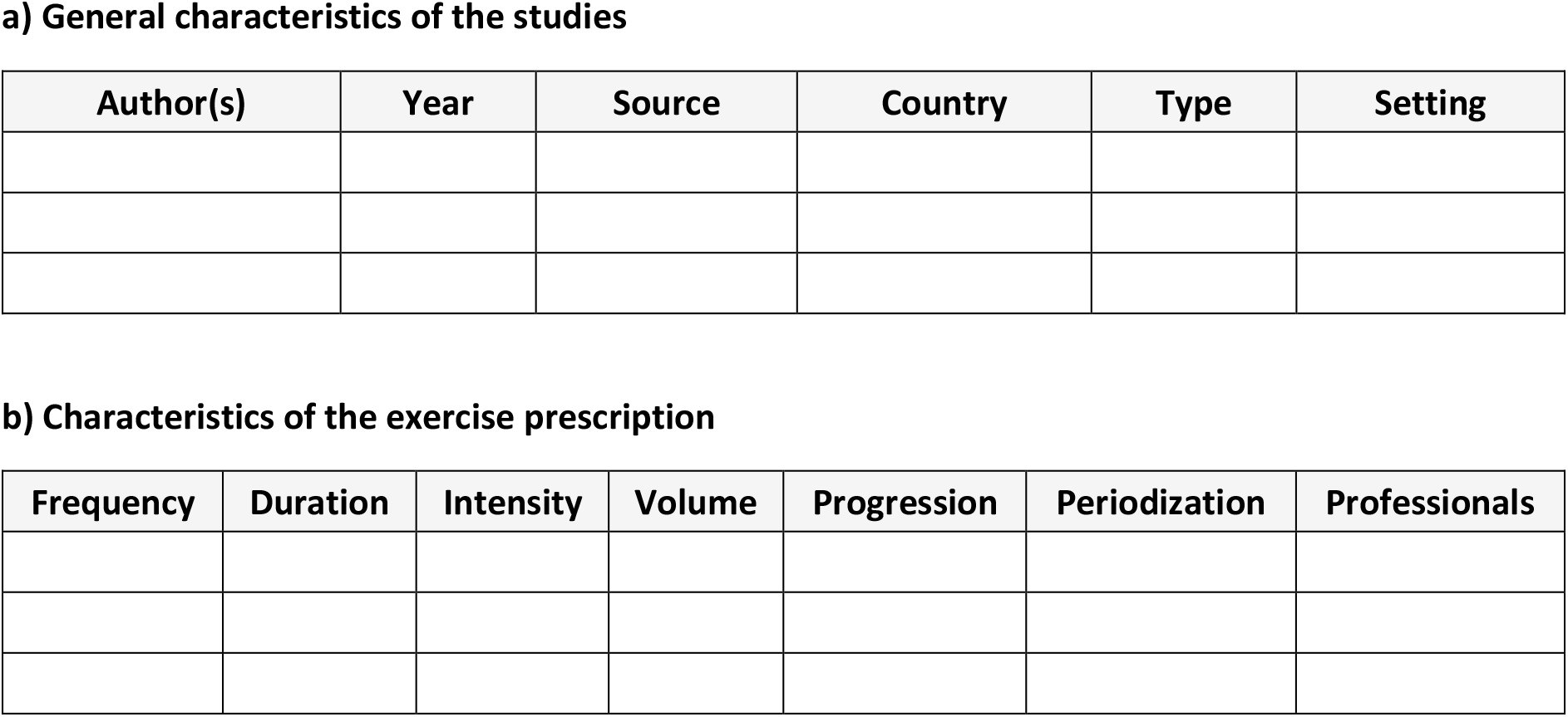

